# Use of artificial intelligence and neural network algorithms to predict arterial blood gas items in trauma victims

**DOI:** 10.1101/2020.03.14.20035584

**Authors:** Milad Shayan, Mohammad Sabouri, Leila Shayan, Shahram Paydar

**Affiliations:** Applied Control & Robotic Research Laboratory (ACRRL), Shiraz University, Shiraz, Iran.; Trauma Research Center, Rajaee (Emtiaz) Trauma Hospital, Shiraz University of Medical Sciences Shiraz, Iran.

**Keywords:** Arterial Blood Gases, Trauma, Neural Network, Prediction

## Abstract

**Background:** Trauma is the third leading cause of death in the world and the first cause of death among people younger than 44 years. In traumatic patients, especially those who are injured early in the day, arterial blood gas (ABG) is considered a golden standard because it can provide physicians with important information such as detecting the extent of internal injury, especially in the lung. However, measuring these gases by laboratory methods is a time-consuming task in addition to the difficulty of sampling the patient. The equipment needed to measure these gases is also expensive, which is why most hospitals do not have this equipment. Therefore, estimating these gases without clinical trials can save the lives of traumatic patients and accelerate their recovery.

**Methods:** In this study, a method based on artificial neural networks for the aim of estimation and prediction of arterial blood gas is presented by collecting information about 2280 traumatic patients. In the proposed method, by training a feed-forward backpropagation neural network (FBPNN), the neural network can only predict the amount of these gases from the patient’s initial information. The proposed method has been implemented in MATLAB software, and the collected data have tested its accuracy, and its results are presented.

**Results:** The results show 87.92% accuracy in predicting arterial blood gas. The predicted arterial blood gases included PH, PCO2, and HCO3, which reported accuracy of 99.06%, 80.27%, and 84.43%, respectively. Therefore, the proposed method has relatively good accuracy in predicting arterial blood gas.

**Conclusions:** Given that this is the first study to predict arterial blood gas using initial patient information(systolic blood pressure (SBP), diastolic blood pressure (DBP), pulse rate (PR), respiratory rate (RR), and age), and based on the results, the proposed method could be a useful tool in assisting hospital and laboratory specialists, to be used.

## I. INTRODUCTION

Trauma is the third leading cause of death in the world and the first cause of death among people younger than 44 years [1,2]. Arterial blood gas (ABG) analysis was proposed and made available by Clark-Caverker nearly 50 years ago [3]. ABG reports essential information to patients [4]. Investigation and monitoring of these gases (ABGs) have a significant impact on the management of intensive care [5,6]. ABG is a gold standard, meaning it is used in the intensive care unit to monitor the respiratory system and the acid-base status of patients with respiratory electrolyte disorders [7]. Measurement of blood pH and oxygen pressure and arterial carbon dioxide determine the patient’s respiratory and metabolic condition. Arterial blood oxygen pressure (paO2) indicates blood oxygenation as well as arterial blood pressure (paCO2), indicating adequate alveolar ventilation. ABG examination, on the other hand, demonstrates the ability of the lungs to absorb oxygen sufficiently and to release adequate carbon dioxide in the blood and lungs, as well as the correct functioning of the kidneys at a pH balance by absorbing or excreting bicarbonate. Sequential measurement of ABG can improve the process of examining lung injury and its progression and analyzing the type of chest injury.

The blood sample needed to test for ABGs can be obtained through superficial arteries or a fixed arterial line to help insert a catheter into the artery. The critical thing about arterial biopsy is that nurses do most of the cases. Nurses should have sufficient information over the interfering factors in the blood sample that can distort the results of the test. Among the essential confounding factors are the time of blood storage and the temperature of blood storage [8,9]. Also, given the techniques available in arterial biopsy in some cases, there is a need for several regular efforts that may lead to painful complications, mainly when performed on the radial artery, including the hematoma, hemorrhage, aneurysm and distal ischemia [10].

ABG measurements are nowadays made using state-of-the-art medical equipment; however, it has many side effects for patients in this process [10]. It is possible to estimate and predict them without arterial biopsy using artificial intelligence and machine learning tools, using primary patient information such as demographics as well as vital signs at the time of admission. There are many statistical methods used for predicting disease progression based on some of the most influential variables, such as discrimination analysis, survival analysis, regression, and artificial neural network [11]. Statistics need basic parameters and assumptions for calculations, and often the data distribution is assumed to be normal and equal variance [12]. Therefore, the results of these methods are highly error-prone, and in many cases, the results are unreliable, so models that do not require these prerequisites are being developed. The most important of these methods can be referred to as the artificial neural network [13]. ANN is one of the machine learning and data mining tools for predicting or classifying data. Data mining is an automated process for extracting patterns that represent knowledge. Data mining simultaneously benefits from several disciplines, such as database technology, artificial intelligence, machine learning, neural networks, statistics, pattern recognition, knowledge-based systems, information retrieval, high-speed computing, and visual data representation [14,15]. One of the most common structures of neural networks is the perceptron structure that uses the feed-forward method. In this structure, the neurons are divided into several layers; in this network, the first layer, the input, the last layer, the output, and the middle layers are called the hidden layers. They can call this architecture the most widely used neural network architecture. Artificial neural networks have been used in many medical fields, including cancer and heart disease, survival prediction, medical image analysis, and others [16,17,18].

The Fabian et al. study was performed in 2005, comparing artificial neural networks and logistic regression for mortality in patients with sepsis. In this study, the area under the rocking curve for logistic regression and artificial neural networks was obtained as 17.75% and 87.8%, respectively. This study demonstrates the accuracy of artificial neural networks relative to logistic regression, respectively [19]. Jutisoni et al. used a neural network to predict heart disease in 2012 [20], using a dataset prepared by Raj Kumar [21]. Sadegh Kara et al. used the artificial neural network to diagnose the optic nerve disease. The final results were classified as healthy and patient [22]. In a case study of 8640 patients, Wang et al. performed a multivariate diagnostic and multivariable diagnostic test for diabetic patients and compared their analysis to the curve-level diagram and achieved higher results [23].

Neural networks have been used for five decades to help physicians diagnose diseases. Neural networks have received particular attention because of their ability to learn in complex issues with the ability to maintain accuracy, even in the absence of some information [24,25], such as in predicting death rate has been used along with statistical methods [26]. Based on previous studies, it can be said that the unique ability of neural networks to differentiate and classify as well as disease detection can be desirable and useful [27,28,29,30,31,32].

Boulain et al., in a study that attempted to predict ABGs from venous blood gases, did not yield favorable results [33]. Honarmand and co-workers have also been able to establish a connection between the blood gases from the earlobe and the ABGs [34]. Concerning smart arterial gas prediction, Wajs et al. have been able to predict arterial blood gas using neural network and arterial gas data obtained from repeated tests over specified time intervals [35].

In this paper, we present an artificial neural-network based approach to predict ABGs based on clinical indicators and vital signs (VSs) that are measured and received at admission. Because of the lack of research on the prediction and estimation of ABG directly using the patient’s baseline indices, the proposed method is essential. The structure of this article is organized as follows. Section 2 presents the structure and concepts of the proposed method. Section 3 presents the simulation results of the proposed method. Section 4 also discusses the results and conclusions of the work.

## II. METHODS AND MATERIAL

The present study was conducted in the Trauma Research Center of Shahid Rajaee (Emtiaz) Hospital in Shiraz, Iran, on data from 2016-2017. The data used in this study included 2280 patients referred to this center. Of these, 374 were female, and 1906 were male. The data used in this study include various variables and indicators that fall into two general categories of clinical and paraclinical signs. Clinical signs include: dyastolic blood pressure (DBP), systolic blood pressure (SBP), pulse rate (PR), respiratory rate (RR), and paraclinical indexes include: PH, PCO2, HCO3. The age parameter has also been added to these indices.

The analysis process in this study is based on the block diagram of Figure 1. In this process, after collecting data at the treatment center and before constructing the artificial neural network and performing the training operation, the data distribution is normalized to improve the training process in the artificial neural network and then to use the formula (1) approx. The data are normalized and are in the interval [0,1]. The data normalization steps were performed in Stata12 and Excel 2013 software.

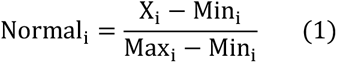

**Figure 1.**
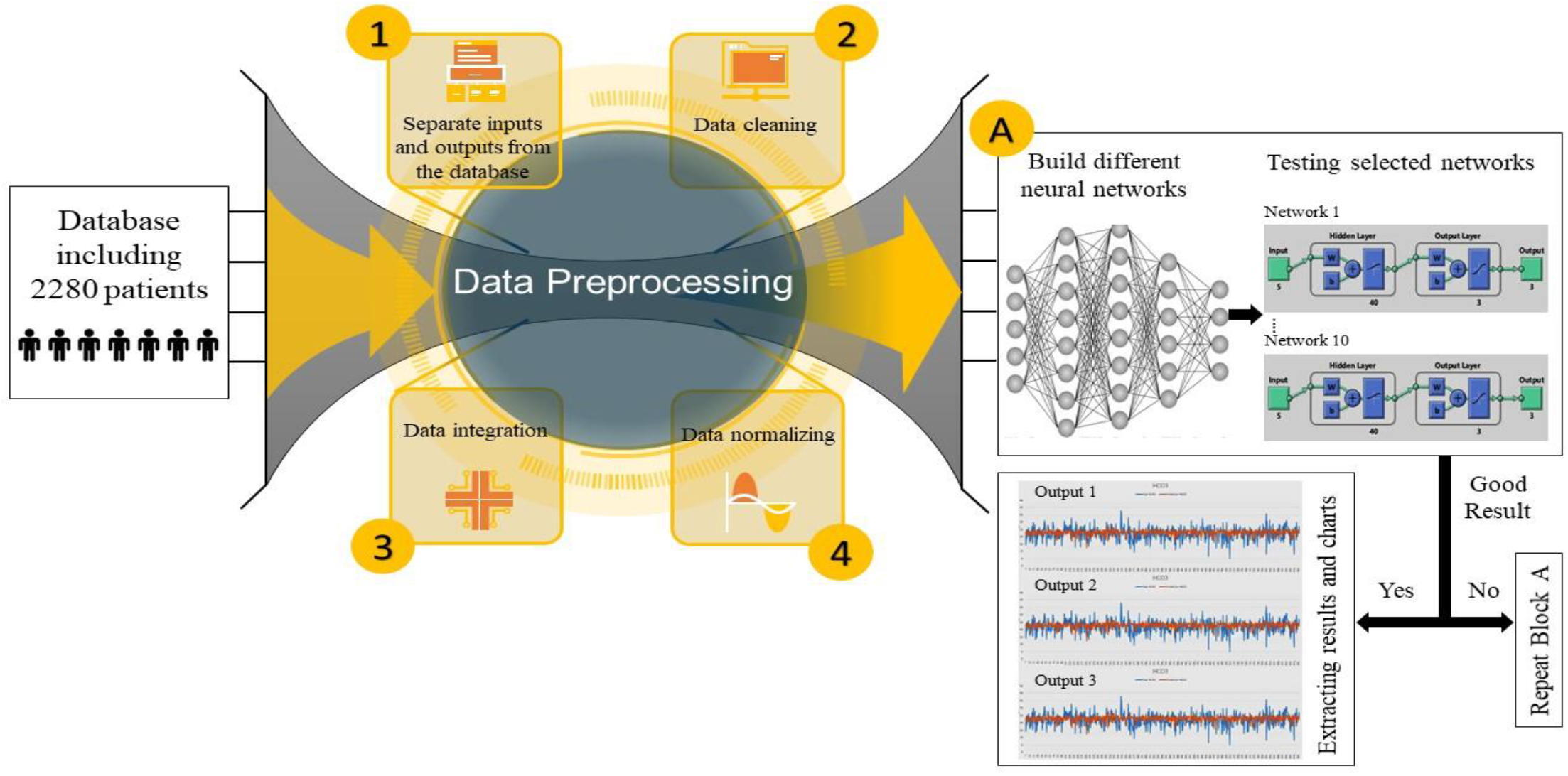
Profile of Study.

Artificial neural networks transmit the knowledge or the law behind the data to the network structure by processing experimental data. That’s why these systems are called smart because they learn general rules based on calculations on numerical data or examples. The structure of these systems have parameters that can be adjusted. The setting of these parameters is to show the system good behavior against external stimuli and information, so-called training of that system. These systems are capable of learning and, through education, gather the knowledge necessary to deal appropriately with a phenomenon and utilize that knowledge when needed [36]. is expressed as follows:

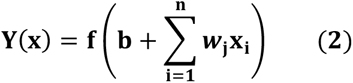

where *Y* is the neural network function with *R^z^* → *R^L^* mapping. *z* and *L* are the number of input and output, respectively. *x_i_* is the *i*th row of neural network input, *w*_*j*_ is the *j*th column of weight matrix, *b* is the network bias vector and *n* is the number of samples.

An artificial neural network is composed of a large number of particular interconnected processing elements called neurons. Each of the neurons in the artificial neural network has a specific weight, each neuron captures the output layer neurons in each layer and transfers its output to each layer neurons after being affected by the activity function. Figure 2 shows the mathematical model of a neuron.

**Figure 2.**
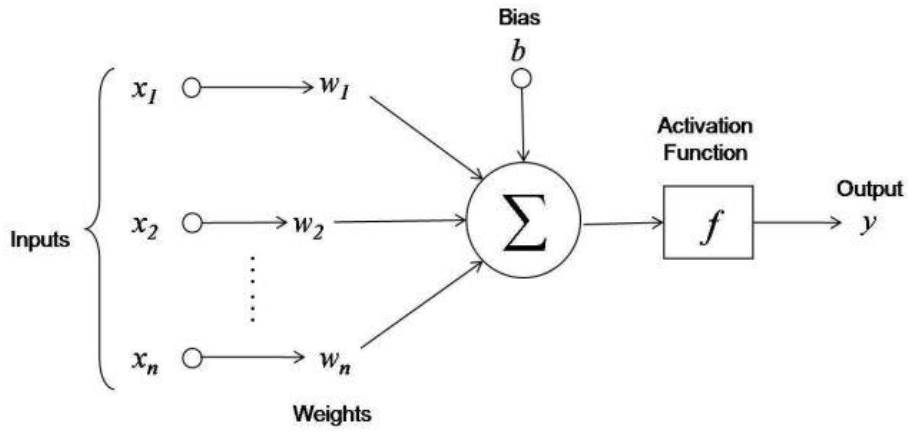
Mathematical structure of a neuron in the neural network.

By expanding Formula 2, which is the structure of a single layer network, a neural network with a hidden layer, an output layer, and an input layer is formed. Formulas 3 and 4 illustrate the designed network mathematics where X is the input data matrix to the network, Wk is the matrix of k-layer weights, bk is the k-layer bias vector, Sq is the number of grid neurons, Y^1^ is the hidden layer output, Y^2^ is the network output, z is the number of inputs, L is the number of outputs and n is the number of input samples, which are given separately in formulas 5 to 8.

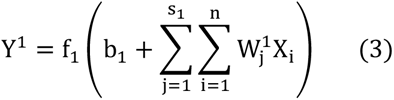

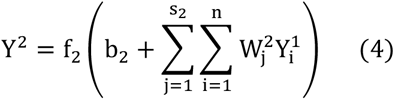

where

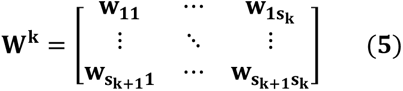

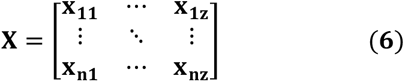

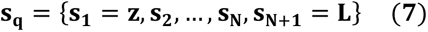

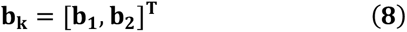

The artificial neural network model used to predict ABG parameters in this study is a multilayer perceptron feedforward neural network, which is one of the most common types of neural networks. In this network, the number of input layer neurons is equal to the number of input vector elements, and the number of output layer neurons is equal to the number of output vector elements [37]. There is no general rule for selecting the number of layers, and most networks use one to three layers and rarely use four-layer or more networks [38,39]. The model structure used in this study has three layers, which include the input layer, the hidden layer (middle), the output layer. The first layer of the network consists of 5 neurons (the number of input data), and its output layer consists of 3 neurons (the number of output data). In general, an accurate estimation of the number of middle layer neurons is complex, and the number of middle layer neurons is a function of the input vector elements. The number of secretory layer neurons is generally obtained experimentally. To enhance the model performance of hidden layer neurons from ten to 100 neurons was selected, the network performance was calculated for the existing structure with a different number of neurons in the hidden layer and finally 40 neurons with the highest performance for the hidden (middle) layer selected. Figure 3 shows the general structure of the neural network used in this study.

**Figure 3.**
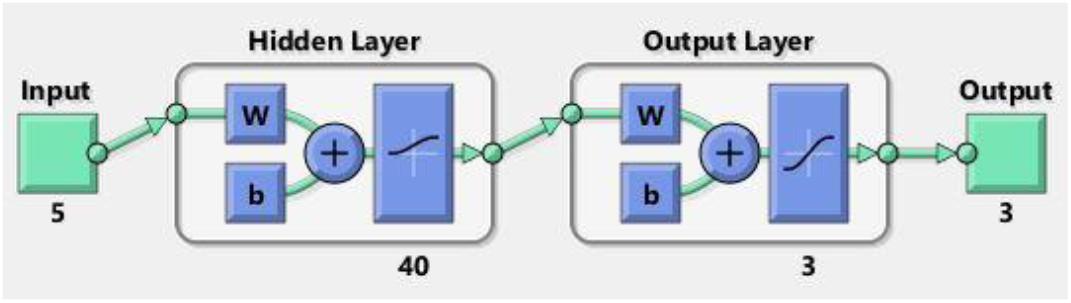
The general structure of the neural network used in this study with 5 input and 3 output and 40 neurons.

Another parameter used in neural networks and its practical implications is the transfer function. The activation function or transfer function determines the relationship between the input and output of a neuron and a network. A network may use different activation functions for different neurons in different or similar layers [40]. Including the most important functions used for thresholding in artificial neural networks, functions such as sigmoid, arctangent, and arc sinus can be noted [41]. Research has shown that networks that use “hyperbolic sigmoidal tangent” or tansig activation functions lead to faster convergence. This function is beneficial for use in neural networks where speed is important [42,43]. Therefore, in this study, logsig and tansig functions have been selected as activation functions and tested.

In general, the network is trained by the rules and data, and by using network learning capabilities, various algorithms are proposed that all try to bring the output generated by the network to the ideal and expected output[44]. The neural network learning mechanism used in this study was based on the concept of descending gradient, which typically uses error propagation law to train this type of network, which has several algorithms. In this study, the backpropagation algorithm is used to reduce the error and calculate the weights in the network. This algorithm uses two sweep paths to minimize the error and approximate the output to the actual value and find the best estimation steps for the parameters. The network continues. This process is called a learning algorithm in artificial neural networks.

An important parameter used in neural networks to evaluate the error in the output of the network is the mean squared error that is equal to the mean square of the difference between the amount of actual arterial blood gas in the body and the value predicted by the network. In the present study, we calculated the relation (9).

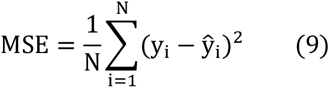

In this formula, N is equal to the number of samples tested, yi is the amount of arterial gases predicted for a sample of i by the proposed method, and y^_i_ is the amount of actual arterial gases sample of i.

In the data analysis process presented with neural-network, these data are divided into two categories of training and final testing, of 2280 tested samples 70% (1596 samples) for network training and 30% (684 samples) for network testing. They are randomized to reduce errors in the process. The data selected for neural network training are obtained by exiting the pre-processing phase through the first layer neurons as inputs and trained according to the set algorithms and various adjustable parameters (Table 1) they take an essential point in the neural-network is to consider the weights and adjustable settings of the neural network. If the weights are selected correctly, the accuracy of the neural network can be increased to an appropriate level [45]. The neural network type used for all ten networks is feed-forward backpropagation with layer number 3.

It is noteworthy that Matlab software and its Neural Network toolbox have been used to build an artificial neural network (suggested model), implement training algorithms, and determine network weights and parameters.

**Table 1.**
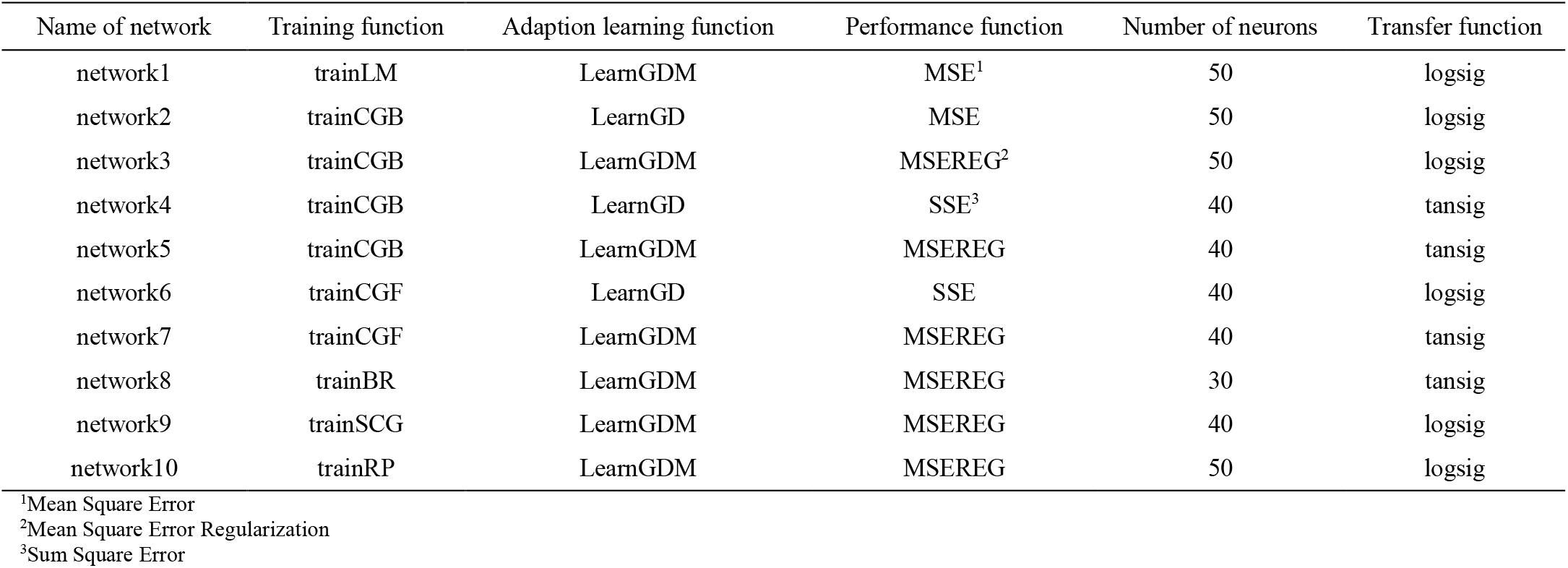
Information on the types of neural networks used in the study.

## III. RESULTS

The current study involved 2280 patients, of which 1596 were directly and 684 indirectly used. The mean age of these patients was 35.93, with a standard deviation of 17.70 years.

The ABG and VS values for all patients on arrival at the registered hospital (raw data) are visible in Table 2. It is important to note that the information and data of patients who participated indirectly in this study were used only for network testing and were not included in the network training process (Table 3).

**Table 2.**
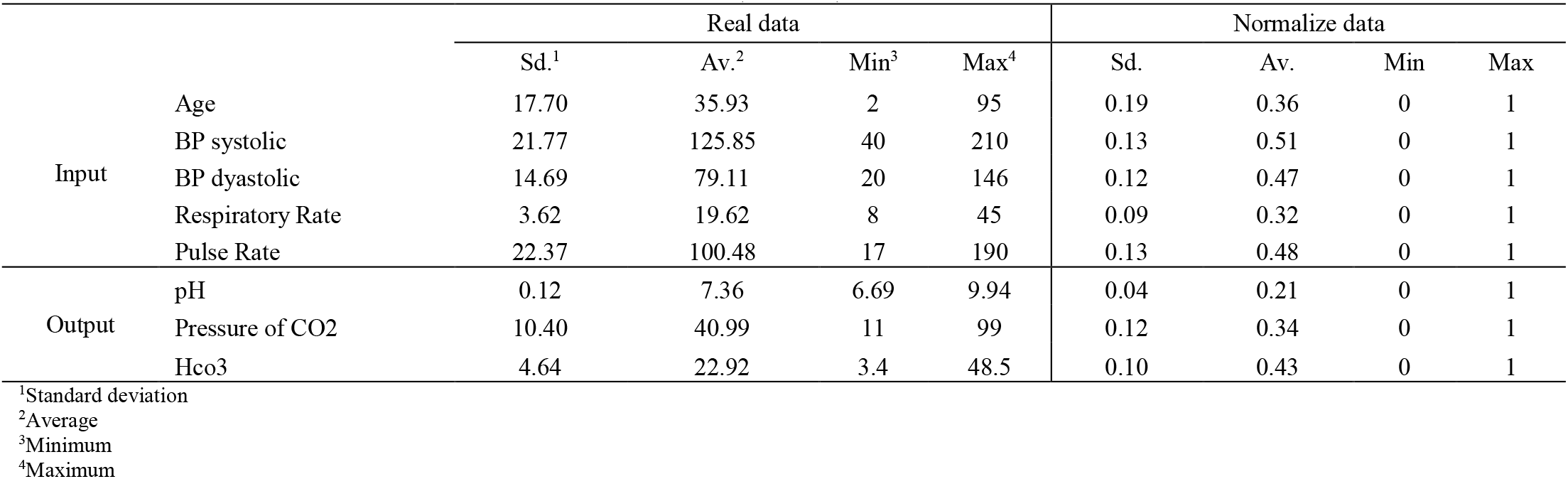
Database information used to train and evaluate neural network (train data)

|  |  | Real data |  |  |  | Normalize data |  |  |  |
| --- | --- | --- | --- | --- | --- | --- | --- | --- | --- |
|  |  | Sd.<sup>1</sup> | Av.<sup>2</sup> | Min<sup>3</sup> | Max<sup>4</sup> | Sd. | Av. | Min | Max |
| Input | Age | 17.70 | 35.93 | 2 | 95 | 0.19 | 0.36 | 0 | 1 |
|  | BP systolic | 21.77 | 125.85 | 40 | 210 | 0.13 | 0.51 | 0 | 1 |
|  | BP dyastolic | 14.69 | 79.11 | 20 | 146 | 0.12 | 0.47 | 0 | 1 |
|  | Respiratory Rate | 3.62 | 19.62 | 8 | 45 | 0.09 | 0.32 | 0 | 1 |
|  | Pulse Rate | 22.37 | 100.48 | 17 | 190 | 0.13 | 0.48 | 0 | 1 |
| Output | pH | 0.12 | 7.36 | 6.69 | 9.94 | 0.04 | 0.21 | 0 | 1 |
|  | Pressure of CO2 | 10.40 | 40.99 | 11 | 99 | 0.12 | 0.34 | 0 | 1 |
|  | Hco3 | 4.64 | 22.92 | 3.4 | 48.5 | 0.10 | 0.43 | 0 | 1 |

<sup>1</sup>Standard deviation
<sup>2</sup>Average
<sup>3</sup>Minimum
<sup>4</sup>Maximum

**Table 3.** Database information used to train and evaluate neural network (test data)

|  |  | Real data |  |  |  | Normalize data |  |  |  |
| --- | --- | --- | --- | --- | --- | --- | --- | --- | --- |
|  |  | Sd.<sup>1</sup> | Av.<sup>2</sup> | Min<sup>3</sup> | Max<sup>4</sup> | Sd. | Av. | Min | Max |
| Input | Age | 18.88 | 38.46 | 4 | 99 | 0.20 | 0.36 | 0 | 1 |
|  | BP systolic | 22.68 | 126.58 | 43 | 223 | 0.13 | 0.46 | 0 | 1 |
|  | BP dyastolic | 16.55 | 79.23 | 23 | 175 | 0.11 | 0.38 | 0 | 1 |
|  | Respiratory Rate | 4.56 | 20.23 | 8 | 45 | 0.12 | 0.33 | 0 | 1 |
|  | Pulse Rate | 22.11 | 97.81 | 15 | 190 | 0.13 | 0.47 | 0 | 1 |
| Output | pH | 0.09 | 7.35 | 6.77 | 7.56 | 0.03 | 0.20 | 0 | 1 |
|  | Pressure of CO2 | 9.93 | 40.46 | 10.3 | 77 | 0.11 | 0.33 | 0 | 1 |
|  | Hco3 | 4.31 | 22.15 | 5.1 | 38.1 | 0.09 | 0.42 | 0 | 1 |

<sup>1</sup>Standard deviation<sup>2</sup>Average<sup>3</sup>Minimum<sup>4</sup>Maximum

After the data were processed, of the 630 possible neural network structures, the data were trained by placing in the ten selected neural networks presented in Table 1, and the results of the network training, test, and error are shown in Table 4. It can be seen from this table that Network 6 has the best result among all the selected networks and has been able to predict the ABG parameters with 87.92% accuracy. According to this table, the network was able to predict the pH, Pco2, and Hco3 parameters with 99%, 80%, and 84% accuracy, respectively.

**Table 4.**
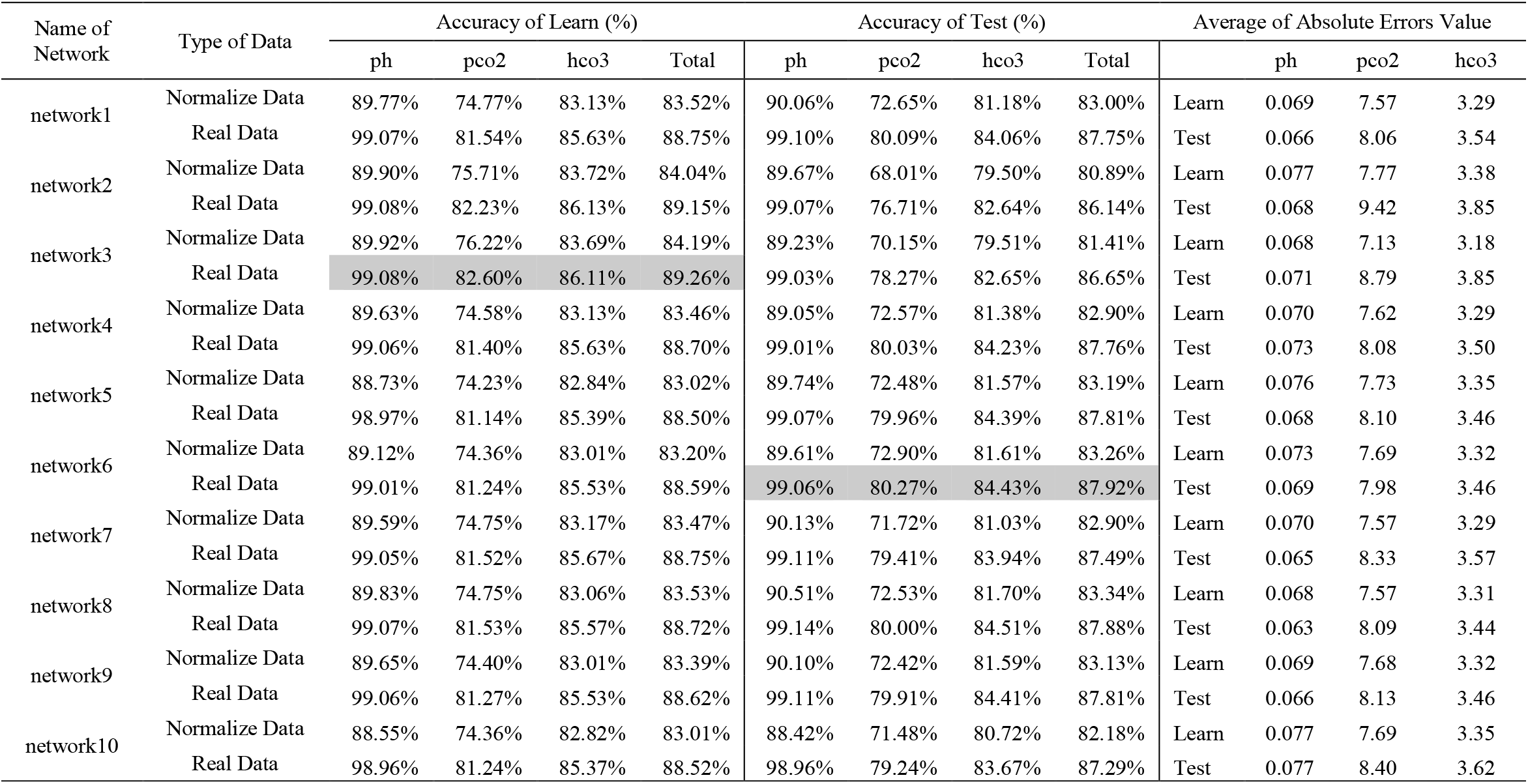
Accuracy of training and test of selected neural networks output with their average error values.

The results of ABG parameter testing using Network6 are proposed in Figure 4 on the data of 684 patients whose networks have never seen their data. Figure 5 also shows the exact output error rate. In Figure 4, the blue graphs are the actual output values, and the red graphs are the values predicted by the neural network. Also shown in Figure 5 is the difference between the output values and the neural network derived from the diagrams in Figure 4.

**Figure 4.**
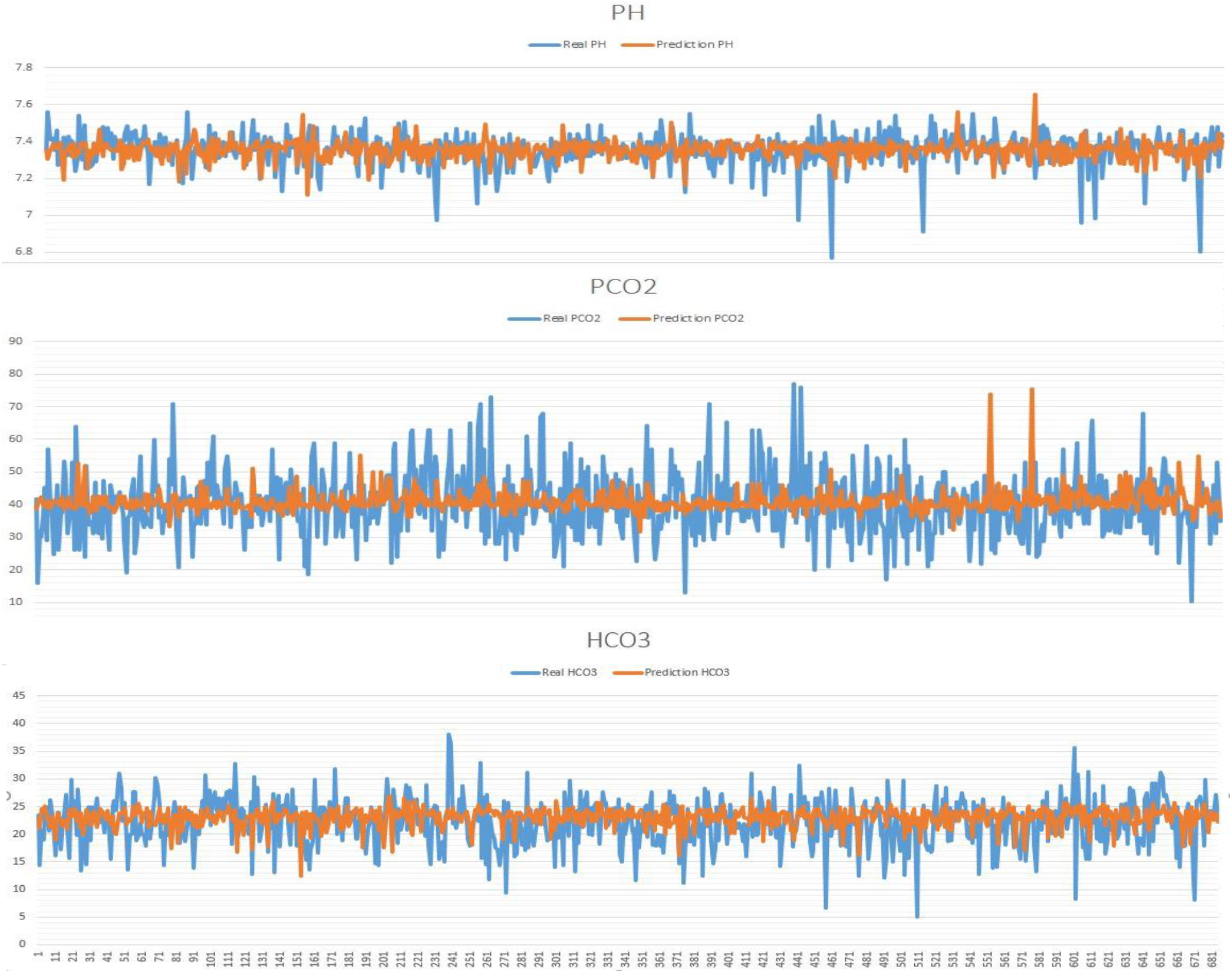
Comparison of arterial blood gas predicted by the neural network with actual arterial blood gas values per test sample.

**Figure 5.**
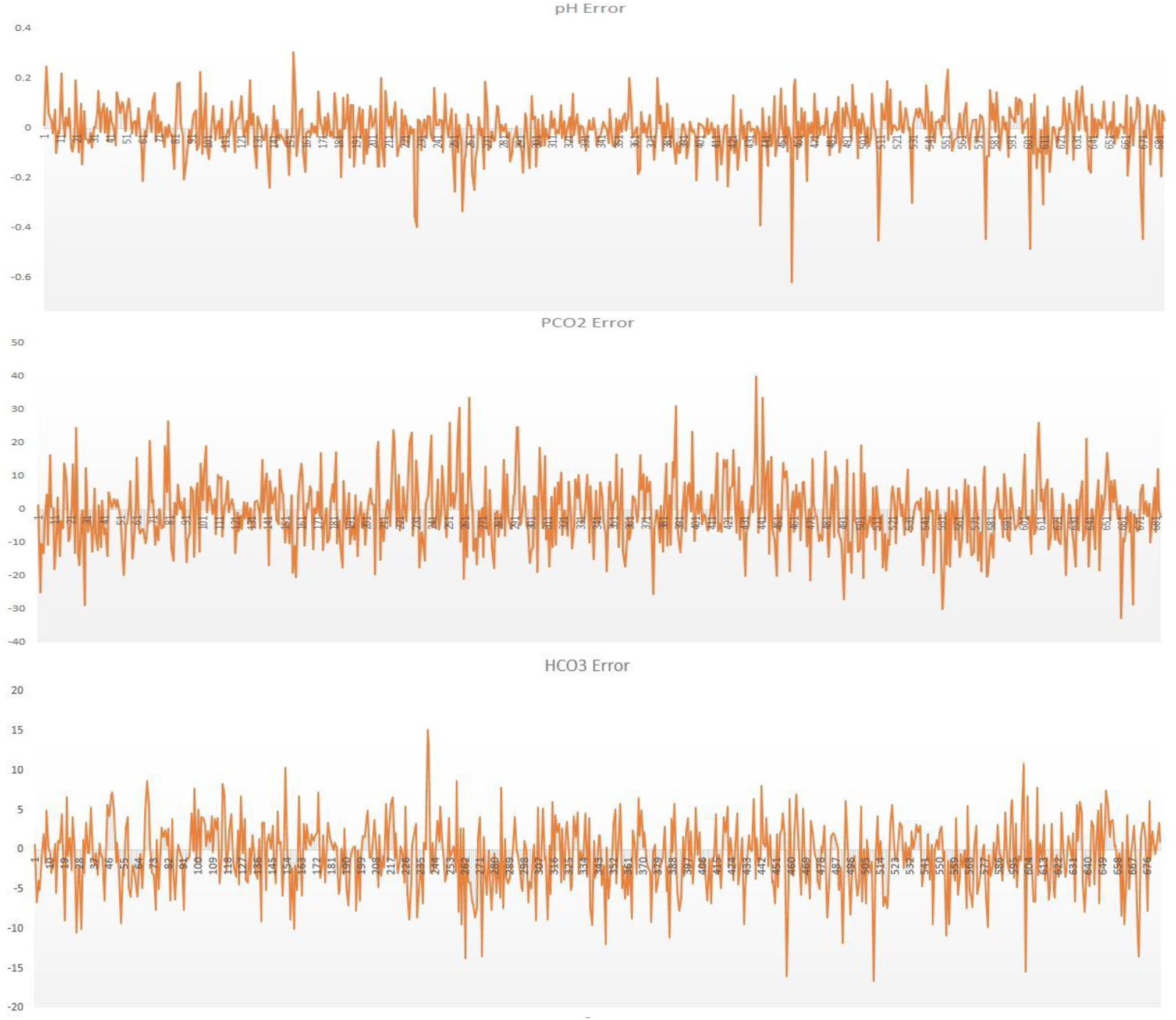
Error in estimating arterial blood gas values per test sample.

## IV. DISCUSSION

The purpose of this study was to obtain an overview of the status of ABG values in traumatic patients so that the medical team can perform the necessary proceedings based on the predicted ABG values without the blood tests and sampling. Knowing the importance of ABG measurements and their impact on the intensive care process in traumatic patients, measuring these gases will be extremely important as physicians calculate them using specialized equipment. Besides, despite the time-consuming process of measuring and calculating these gases, as well as the difficulties involved in the process of blood sampling from patients and the cost of measuring and laboratory equipment, ABG is calculated for conditions that ABG is not available for managing trauma victims. In this regard, using the tools and facilities that make this complicated process more comfortable and improve the calculation process can enhance the quality of treatment and reduce its associated costs. Therefore, using artificial intelligence tools to predict the amount of these gases, in addition to improving the process and reducing costs, plays a vital role in the chances of trauma survivors being able to improve the quality and speed up the process of effective early action. While artificial intelligence and machine learning tools have been used in many medical fields, including cancer and heart disease, survival prediction, medical image analysis, and others [12,13,14], predicting and estimating the amount of ABG by these smart methods there are very few studies in the world. In the present study, based on an artificial neural network, five features were used as inputs, and an accuracy of about 87.92% was obtained.

There have been numerous studies to date on the relationship between arterial blood gas and venous blood gas through statistical methods and analyses. Still, it can be stated that no studies have been performed to estimate and predict these gases from clinical and preliminary patient information by artificial neural networks that have not been directly sampled. In a 2008 study, Honarmand and colleagues were able to find a link between earlobe blood gases and ABGs. They performed their tests on 60 patients admitted to the intensive care unit treated with mechanical ventilation and took blood samples from the radial artery and arterialized earlobe at the same time. Finally, regression equations were used predicting ABG levels and investigating the correlation between pH, PCO2, PO2, BE, and HCO3-values of blood obtained from earlobe and arterial blood have shown that there is a significant relationship between pH, PCO2, BE and HCO3 have been the case between earlobe and arterial blood samples in patients with normal blood pressure who have mechanical ventilation. While the authors of the study stated that they could use blood samples taken from the earlobe instead of the arterial blood sample for testing and calculating ABG, but because of the low statistical population in this study and the percentages expressed, the accuracy of this method can be argued with certainty. Also, like other methods for measuring and calculating ABG, it requires blood sampling and the use of advanced laboratory apparatus [34].

In another study by Boulain et al., 2016, statistical methods were used to predict ABGs from intravenous blood gases. In this study, 590 patients with acute circulatory failure were enrolled, and their simultaneous venous and arterial blood samples were measured at specified intervals over 24 hours. Although in this study the relative results of the relationship between venous and arterial gases and the clinical information of patients were obtained, the predicted outputs were of poor accuracy, according to the conclusions made by the authors of this study. Alongside this problem, the research method chosen by the researchers in this study is a very time consuming and costly one [33].

Wajs et al. (2016) researched smart blood gas prediction. They have been able to predict ABGs using neural network and ABG data obtained from repeated tests over a specified period. The prediction made in this study is based on the calculation of a time series dependent on the patient dynamics model obtained from data collected from a patient. The neural network used is a dynamic network that is retrained by receiving any data, which creates a computational time problem. It also seems that the model presented in this study is not comprehensive because of its dependence on the patient’s condition and requires the collection of preliminary data to construct it [35].

In the present study, an artificial neural network with a hidden layer was used to predict and estimate ABGs in patients with an average error of 12.08% and a mean unit of 0.069 (0.94% error), respectively for Ph, 7.9844 (19.73% error) for Pco2 and 3.4561 (15.57% error) for Hco3. However, the proposed method does not require blood sampling of patients. It performs prediction using only clinical information of patients, and obtaining data including age, Bps, Bpd, RR, PR, and neural network calculates the amounts of gases. The proposed method also calculates ABGs without the need for prior data, so it can predict when the patient arrives at the treatment center in the shortest possible time. The results of this study showed that the model and method used for estimating ABGs without the need for sampling work. Therefore, it can be stated that using the presented training model, the use of laboratory devices, and the time required to calculate and perform the relevant experiments to calculate these gases is minimized. Besides, because of the reduced time of measurement and estimation of these gases, early treatment can be best performed on patients, especially trauma patients whose timing and appropriate therapies are of the utmost importance.

## V. CONCLUSION

In this study, we were able to predict arterial blood gas items in traumatic patients using artificial intelligence and neural network. The neural network structure used in this study is a multilayer perceptron feedforward neural network with three hidden layers. Using data from 2280 patients including clinical indicators: diastolic blood pressure (BPD), systolic blood pressure (BPS), pulse rate (PR), respiratory rate (RR), and paraclinical indices: PH, PCO2, HCO3 as well as patients’ age was able to predict ABGs of 684 patients (test data) with accuracy of 87.92% by training 1596 patients (training data). This is very important given the difficulties in measuring ABGs in the conventional and time-consuming way. It is hoped to increase this accuracy by combining the neural network with other artificial intelligence methods.

## Data Availability

The data used in this study were collected from patients of Shahid Rajaee (Emtiaz) Hospital in Shiraz. This data is currently available to the center.

## Abbreviations

ABG: Arterial Blood Gas
BPD: Blood Pressure Diastolic
BPS: Blood Pressure Systolic
RR: Respiratory Rate
PR: Pulse Rate
VS: Vital Sign

## Acknowledgments

The authors would like to thank the members of Shiraz Trauma Research Center of Shahid Rajaee (Emtiaz) Hospital members, especially Dr. Shahram Bolandparvaz, head of Trauma Research Center, for providing this research opportunity, and also Dr. HamidReza Abbasi, deputy of Trauma Research Center for his cooperation and supporting in this research.

## Authors’ contributions

MSH and MS contributed to the study conception, implementation of the study, design, interpretation and critical was involved in drafting of the manuscript. SHP and LSH contributed to the conception and design of data and revising the manuscript. All authors have read and approved the manuscript, and ensure that this is the case.

## Availability of data and materials

The data that support the findings of the present study are available from the Rajaee (Emtiaz) Hospital and are not publicly available. The anonymized dataset used for the present research is however available from the corresponding author on reasonable request and with permission of both Trauma Research Center and Rajaee (Emtiaz) Hospital of Shiraz.

## Ethics approval and consent to participate

This study was conducted according to the principles expressed in the Trauma Research Center of Shahid Rajaee (Emtiaz) Hospital of Shiraz and approved by the local ethics committee of Shiraz University of Medical Sciences by the code IR.SUMS.REC.1397.719. Based on the approval of Ethics Committee, all informations were collected only by patient’s code and their identity was not disclosed. Patient’s information was in private.

## Consent for publication

Not applicable.

## Author details

^1^Applied Control & Robotic Research Laboratory (ACRRL), Shiraz University, Shiraz, Iran. ^2^Applied Control & Robotic Research Laboratory (ACRRL), Shiraz University, Shiraz, Iran. ^3^Trauma Research Center, Rajaee (Emtiaz) Trauma Hospital, Shiraz University of Medical Sciences Shiraz, Iran. ^4^Trauma Research Center, Rajaee (Emtiaz) Trauma Hospital, Shiraz University of Medical Sciences Shiraz, Iran.

